# Childhood asthma outcomes during the COVID-19 pandemic: Findings from the PeARL multi-national cohort

**DOI:** 10.1101/2020.10.27.20219436

**Authors:** Nikolaos G. Papadopoulos, Alexander G. Mathioudakis, Adnan Custovic, Antoine Deschildre, Wanda Phipatanakul, Gary Wong, Paraskevi Xepapadaki, Rola Abou-Taam, Ioana Agache, Jose A. Castro-Rodriguez, Zhimin Chen, Pierrick Cros, Jean-Christophe Dubus, Zeinab Awad El-Sayed, Rasha El-Owaidy, Wojciech Feleszko, Vincenzo Fierro, Alessandro Fiocchi, Luis Garcia-Marcos, Anne Goh, Elham M. Hossny, Yunuen R. Huerta Villalobos, Tuomas Jartti, Pascal Le Roux, Julia Levina, Aida Inés López García, Ángel Mazón Ramos, Mário Morais-Almeida, Clare Murray, Karthik Nagaraju, Major K Nagaraju, Elsy Maureen Navarrete Rodriguez, Leyla Namazova-Baranova, Antonio Nieto Garcia, Cesar Fireth Pozo Beltrán, Thanaporn Ratchataswan, Daniela Rivero Yeverino, Eréndira Rodríguez Zagal, Cyril E Schweitzer, Marleena Tulkki, Katarzyna Wasilczuk, Dan Xu, PeARL collaborators, on behalf of the PeARL Think Tank

## Abstract

**Importance:** Importance: The interplay between COVID-19 pandemic and asthma in children is still unclear.

**Objective:** We evaluated the impact of COVID-19 pandemic on childhood asthma outcomes.

**Design:** The PeARL multinational cohort included children with asthma and non-asthmatic controls recruited during the COVID-19 pandemic and compared current disease activity with data available from the previous year.

**Setting:** Pediatric outpatient clinics.

**Participants:** The study included 1,054 children with asthma and 505 non-asthmatic controls, aged between 4-18 years, from 25 pediatric departments, from 15 countries globally.

**Exposures:** COVID-19 pandemic first wave, starting from the date of the first fatality in the respective country.

**Main outcomes and measures:** We assessed the pandemic’s impact on the frequency of respiratory infections, emergency presentations and hospital admissions in asthmatic versus non-asthmatic participants, controlling for confounding factors including the pandemic’s duration and the frequency of such acute events during 2019. Using paired analyses, we evaluated the impact of the pandemic on the annualized frequency of asthma attacks and the previously mentioned acute events, asthma control, and pulmonary function in children with asthma, compared to their baseline disease activity, during the preceding year.

**Results:** During the pandemic, children with asthma experienced fewer upper respiratory tract infections, episodes of pyrexia, emergency visits, hospital admissions, asthma attacks and hospitalizations due to asthma, in comparison to the preceding year. Sixty-six percent of asthmatic children had improved asthma control while in 33% the improvement exceeded the minimally clinically important difference. Pre-bronchodilatation FEV1 and peak expiratory flow rate were also improved during the pandemic.

When compared to non-asthmatic controls, children with asthma were not found to be at increased risk of LRTIs, episodes of pyrexia, emergency visits or hospitalizations during the pandemic. However, an increased risk of URTIs emerged.

**Conclusions and relevance:** Childhood asthma outcomes, including control, were improved during the first wave of the COVID-19 pandemic, probably because of reduced exposure to asthma triggers and increased treatment adherence. The decreased frequency of acute episodes does not support the notion that childhood asthma may be a risk factor for COVID-19. Furthermore, the potential for improving childhood asthma outcomes through environmental control becomes apparent.

**Key Points:** *Question:* What was the impact of COVID-19 pandemic on childhood asthma outcomes?

*Findings:* During the first wave of the pandemic, children with asthma have experienced improved outcomes, as evidenced by fewer asthma attachks, hospitalizations, improved scores in validated asthma control measures and improved pulmonary function.

*Meaning:* This is the first study to show a positive impact of COVID-19 pandemic on childhood asthma activity. This is probably the result of reduced exposure to asthma triggers and increased treatment adherence. The decreased frequency of acute episodes does not support the hypothesis that childhood asthma may be a risk factor for COVID-19.

## INTRODUCTION

A series of studies have demonstrated that the morbidity from Coronavirus Disease 2019 (COVID-19) is lower in children compared to adults^1,2,3^. Risk factors for severe disease include older age, male sex, chronic respiratory diseases, diabetes, coronary artery disease, obesity and ethnicity (black, Asian and mixed)^4,5,6^. Chronic respiratory diseases are among these high-risk pre-existing conditions and asthma represents the majority of such patients^4,5,7^. Initial clinical reports did not identify asthma to be over-represented among COVID-19 patients^8^, but severe, uncontrolled asthma is included in most guidance documents among conditions which may increase the risk for severe COVID-19^9^. Analysis in more than 17 million adults and ∼11,000 COVID-19 related deaths identified severe asthma (defined as asthma with recent oral corticosteroid use) as a significant associate of COVID-19 death^6^. It is however unclear what is the impact of asthma on the risk of SARS-CoV-2 infection and severe COVID-19 in children, and what is the impact of COVID-19 pandemic on asthma-related outcomes in children. Survey data among pediatric asthma specialists suggest that there is no apparent increase in asthma-related morbidity in children with asthma^10^; it is even possible that due to increased adherence and reduced exposures due to confinement, such children may have improved outcomes^10^. Furthermore, it is also possible that allergic sensitization can have some protective effect against COVID-19^11^. However, this needs to be further explored^12,13^.

Recent systematic reviews have concluded that there are limited data on pediatric asthma as a risk factor for SARS-CoV-2 infection or COVID severity and called for data^2,14,15^. For the above reasons, Pediatric Asthma in Real Life (PeARL), a think tank initiated by the Respiratory Effectiveness Group (REG), comprising of pediatric asthma experts globally^16^, opted to evaluate the interplay between childhood asthma and COVID-19 infection in a multi-national cohort of children with asthma and non-asthmatic controls. We aimed at assessing asthma activity (asthma control, respiratory infections, asthma attacks, lung function) during the first months of the COVID-19 pandemic and exploring whether children with asthma had excess morbidity during this period, in comparison to prerecorded historical data.

The primary study objective was to assess differences in the impact of the pandemic on the frequency of upper (URTIs), lower respiratory tract infections (LRTIs), episodes of fever, emergency visits and hospital admissions between asthmatic and non-asthmatic children. Secondary objectives to evaluate were: (i) the impact of the pandemic on the frequency of asthma attacks, as well as the previously mentioned events among asthmatic children, compared to the baseline frequency of these events, during the preceding year (2019); (ii) the impact of the pandemic on disease control, evaluated using validated asthma clinical questionnaires, among children with asthma, compared to the baseline asthma control, during the preceding year (2019).

## METHODS

### Study design, setting and participants

The impact of the COVID-19 pandemic on disease activity in children with asthma was evaluated in data collected as part of a multinational audit. Ethics review was not required for this audit, in most participating countries. When required, an ethics approval was acquired. The cross-sectional case-control study was designed and managed by the PeARL Steering Committee, was conducted in accordance with the Good Clinical Practice Guidelines and reported following the Strengthening the reporting of observational studies in epidemiology (STROBE) statement^17^. Participating centers were identified among members and collaborators of the PeARL think-tank.

### Eligibility criteria, sources and methods of selection of participants

Eligible subjects were children aged between 4 and 18 years, diagnosed with asthma and monitored in one of the participating asthma clinics. Non-asthmatic controls included children of the same age range, monitored at the same healthcare setting for a non-respiratory condition, who did not clinically suffer from asthma. Children with other chronic respiratory conditions, diabetes, hypertension, immunodeficiency, or any other chronic disease deemed by the responsible clinician that could impact the main outcomes of this study, were excluded. Each participating center was asked to contribute data from at least 45 participants, including twice as many children with asthma, compared to controls.

### Definition of variables

For the purposes of this study, the onset of the pandemic in each participating center was defined as the date of the first fatality due to COVID-19 in the respective country. For each participant, the duration of the pandemic period was defined as the period between the onset of the pandemic and the captured clinical visit. Historical data (before the pandemic) were captured during 2019 and the duration of observation before the onset of the pandemic was one year, for all participants.

We collected basic demographic data for all participants and data on the main outcomes of interest. The frequency of acute events (URTIs, LRTIs, asthma attacks, episodes of fever, emergency visits, for any reason, and hospital admissions, for any reason) during and before the pandemic were reported as recalled by the participants or their parents. Asthma attacks were defined as the need for systemic glucocorticoids administration for at least 48 hours or an emergency visit or a hospitalization for asthma. Need for additional treatment was defined as any treatment escalation, including increased use of short-acting bronchodilators, increased dose of inhaled corticosteroids or the administration of systemic corticosteroid courses. Asthma activity during and before the pandemic was measured using validated disease control questionnaires (asthma control test: ACT^18^, childhood asthma control test: C-ACT^19^, asthma control questionnaire: ACQ^20^, or composite asthma severity index: CASI^21^). For each participant with asthma, at least one questionnaire was administered at least 2 months after the onset of the pandemic in their country and at least once during 2019 (the same questionnaire in both occasions). Physiological measurements, including forced expiratory value in 1 second (FEV1), before or after bronchodilation and the peak expiratory flow rate (PFR), during and before the pandemic, were also captured when available.

### Statistical analysis

Statistical analyses were performed using R statistical software (version 3.4.3 or newer; R Foundation for Statistical Computing; Vienna; Austria). We did not perform power calculations, as this was an exploratory analysis. Shapiro-Wilk test was used to evaluate normality of the distribution of continuous data. Student’s t-test and Mann-Whitney test were used for comparing means of normally or non-normally distributed variables, respectively. Chi-squared test was used for comparing dichotomous data. The level of significance was set at 0.05 for all analyses.

We used a generalized linear model for evaluating the impact of the pandemic on the frequency of acute events in asthmatic versus non-asthmatic participants. We assumed a negative binomial distribution, with the number of events during the pandemic as the response variable and the natural logarithm of the duration of the pandemic for each participant as an offset variable. At first, based on established theory, we considered as potential covariates the age, gender, and the number of acute events each participant experienced during 2019. In an additional model we also accounted for the race (if it was available, according to the participating countries’ ethics regulations), passive smoking history (cigarettes smoked per day by the parents), concomitant allergic rhinitis, IgE sensitization, food allergy and history of flu vaccination during the preceding year.

For evaluating the impact of the pandemic on the frequency of each acute event in asthmatic and non-asthmatic children, we extrapolated an annualized event count based on the number of observed events during the pandemic (365 * events / pandemic duration [in days]). We then compared this with the actual number of acute events observed during 2019 using the Wilcoxon Signed-Rank test. In addition, we present the number of participants who had at least one event during the pandemic and during the preceding year.

Wilcoxon Signed-Rank test was also used to assess the impact of the pandemic on asthma control. As previously described, different standardized measures of asthma control were used in each center, depending on the measures that were evaluated during 2019, to facilitate comparisons. Firstly, we compared asthma control during versus before the pandemic in subgroups of participants, depending on the available asthma control tools. Next, we used Z-scores to estimate standardized differences by dividing the differences in the values during versus before the pandemic, with the standard deviation of all differences, for every test. In a sensitivity analysis, we only included subjects with a previous asthma control assessment between March-June 2019, during the same period as the pandemic, to account for seasonal variation.

Paired t-test was used to compare pulmonary function measurements during versus before the pandemic. As previously, we conducted a sensitivity analysis only including subjects with a previous pulmonary function test, between March-June, 2019.

Finally, in a subgroup analysis we repeated all previously described analyses only including data from participating centers that are in countries that were more severely hit by the pandemic (>200 deaths per million of inhabitants by 13/07/2020).

## RESULTS

### Participants and descriptive data

The study included 1,054 children with asthma and 505 control subjects, from 25 pediatric departments from 15 countries globally (supplementary table 1). The baseline characteristics of the participants were generally balanced between the groups with a few, anticipated exceptions (Table 1-2). A higher proportion of males (62.8% versus 51.9%) was observed in the asthma group compared to the controls, which is consistent with epidemiological characteristics of the disease in this age group. Allergic diseases such as allergic rhinitis (79.4% vs 39.4%) and food allergy (22.8% vs 15.6%) were also more prevalent in the asthma group. A higher proportion of children with asthma had a confirmed IgE sensitization and were vaccinated for the flu in the preceding year. Finally, children with asthma suffered a higher number of episodes of URTIs, LRTIs, emergency visits or hospital admissions during 2019, compared to the control group (Table 3).

**Table 1:**
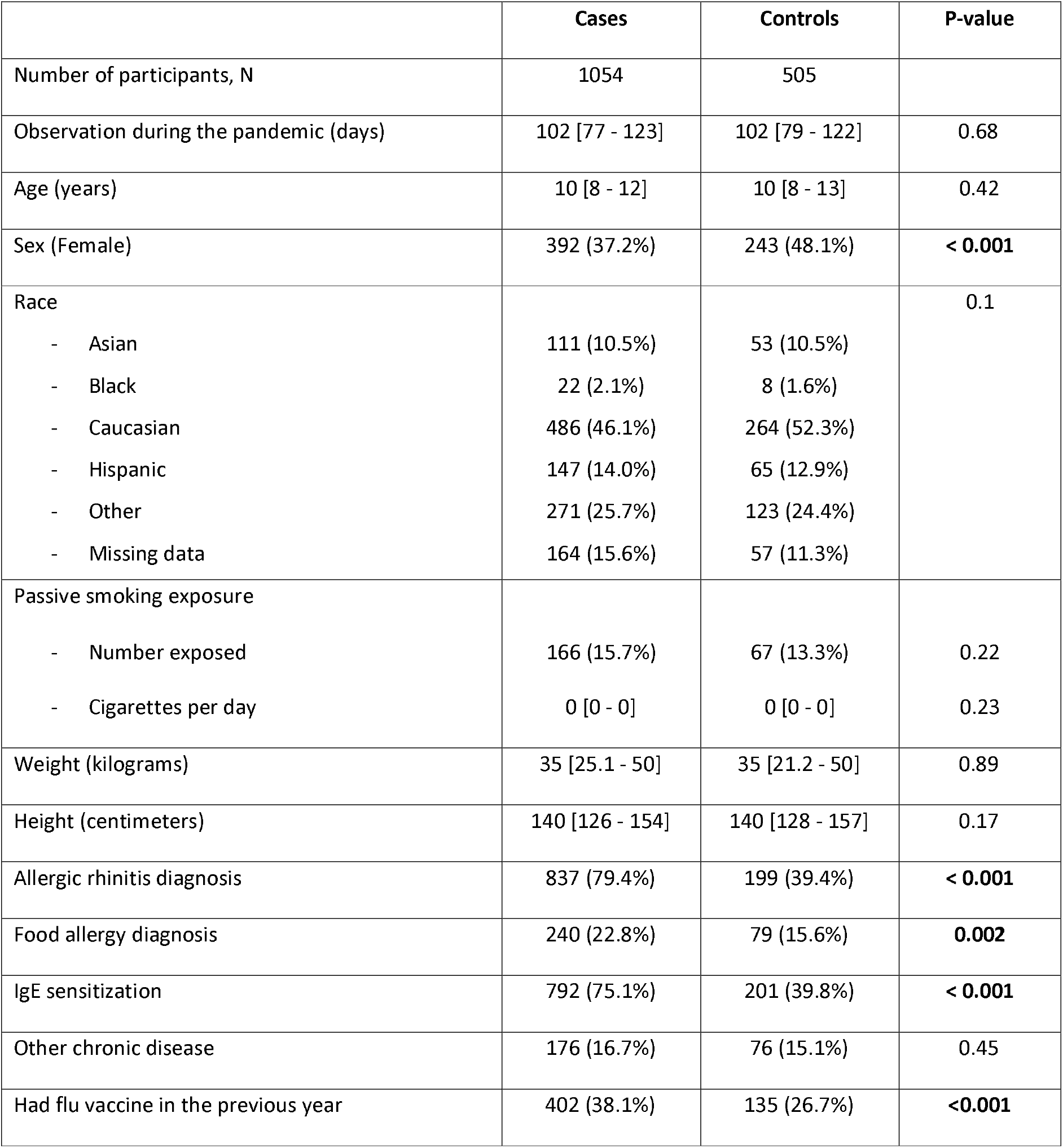
Main characteristics of the participants. Continuous data are presented as median [percentiles 25-75], total number of participants with available data for this variable. and dichotomous data as number of events/ total numbers (percentage).

|  | Cases | Controls | P-value |
| --- | --- | --- | --- |
| Number of participants, N | 1054 | 505 |  |
| Observation during the pandemic (days) | 102 [77 - 123] | 102 [79 - 122] | 0.68 |
| Age (years) | 10 [8 - 12] | 10 [8 - 13] | 0.42 |
| Sex (Female) | 392 (37.2%) | 243 (48.1%) | **< 0.001** |
| Race |  |  | 0.1 |
| - Asian | 111 (10.5%) | 53 (10.5%) |  |
| - Black | 22 (2.1%) | 8 (1.6%) |  |
| - Caucasian | 486 (46.1%) | 264 (52.3%) |  |
| - Hispanic | 147 (14.0%) | 65 (12.9%) |  |
| - Other | 271 (25.7%) | 123 (24.4%) |  |
| - Missing data | 164 (15.6%) | 57 (11.3%) |  |
| Passive smoking exposure |  |  |  |
| - Number exposed | 166 (15.7%) | 67 (13.3%) | 0.22 |
| - Cigarettes per day | 0 [0 - 0] | 0 [0 - 0] | 0.23 |
| Weight (kilograms) | 35 [25.1 - 50] | 35 [21.2 - 50] | 0.89 |
| Height (centimeters) | 140 [126 - 154] | 140 [128 - 157] | 0.17 |
| Allergic rhinitis diagnosis | 837 (79.4%) | 199 (39.4%) | **< 0.001** |
| Food allergy diagnosis | 240 (22.8%) | 79 (15.6%) | **0.002** |
| IgE sensitization | 792 (75.1%) | 201 (39.8%) | **< 0.001** |
| Other chronic disease | 176 (16.7%) | 76 (15.1%) | 0.45 |
| Had flu vaccine in the previous year | 402 (38.1%) | 135 (26.7%) | **<0.001** |

**Table 2:** Asthma severity and asthma treatment adherence in the asthma group.

|  |  |
| --- | --- |
| **Asthma severity:** |  |
| - Intermittent | 296 (28.1%) |
| - Mild persistent | 351 (33.3%) |
| - Moderate persistent | 291 (27.6%) |
| - Severe persistent | 111 (10.5%) |
| - Unknown | 5 (0.5%) |
| **Treatment adherence:** |  |
| - Always as prescribed | 551 (52.3%) |
| - Sometimes not taken | 282 (26.8%) |
| - Often not taken | 41 (3.9%) |
| - Never taken | 106 (10.1%) |
| - Unknown | 74 (7.0%) |

**Table 3:** Acute events observed during 2019 and during the pandemic. Presented as median [percentiles 25-75], and the percentage of participants experiencing at least one event in the respective observation period. Between group differences assessed using generalized linear models. The first model considers the age, gender and historical number of acute events as covariates, while the second model also accounts for the race, passive smoking history, concomitant allergic rhinitis, IgE sensitization, food allergy and history of flu vaccination during the preceding year. <u>* Events per 100 participants per year.</u>

| Acute events | Children with asthma |  |  | Controls |  |  |
| --- | --- | --- | --- | --- | --- | --- |
|  | 2019 | Pandemic | Frequency<br>change during<br>pandemic | 2019 | Pandemic | Frequency<br>change during<br>pandemic |
| URTI | 2 [1 - 4],<br>81.6% | 0 [0 - 1],<br>30.9% | *p<0.001*<br>*73 fewer\** | 2 [0 - 3],<br>71.3% | 0 [0 - 1],<br>26.1% | *p<0.001*<br>*35 fewer\** |
| LRTI | 0 [0 - 1],<br>28.8% | 0 [0 - 0],<br>12.1% | *p=0.12* | 0 [0 - 0],<br>16.8% | 0 [0 - 0],<br>9.1% | *p<0.001*<br>*18 more\** |
| Pyrexia | 0 [0 - 2],<br>52.4% | 0 [0 - 0],<br>17.2% | *p<0.001*<br>*24 fewer\** | 0 [0 - 2],<br>51.5% | 0 [0 - 0],<br>15.8% | *p<0.001*<br>*20 fewer\** |
| Emergency visit | 0 [0 - 1],<br>34.4% | 0 [0 - 0],<br>11.0% | *p<0.001*<br>*9 fewer\** | 0 [0 - 0],<br>20.0% | 0 [0 - 0],<br>8.5% | *p=0.79* |
| Hospital<br>admission | 0 [0 - 0],<br>13.1% | 0 [0 - 0],<br>2.0% | *p<0.001*<br>*17 fewer\** | 0 [0 - 0],<br>5.5% | 0 [0 - 0],<br>1.8% | *p=0.29* |
| Need for<br>additional<br>treatment | 2 [0 - 4],<br>74.0% | 0 [0 - 1],<br>29.5% | *<0.001*<br>*165 more\** |  |  |  |
| Acute asthma | 0 [0 - 1],<br>40.7% | 0 [0 - 0],<br>9.6% | *p<0.001*<br>*31 fewer\** |  |  |  |
| Hospitalization<br>for acute<br>asthma | 0 [0 - 0],<br>9.8% | 0 [0 - 0],<br>1.2% | *p<0.001*<br>*9 fewer\** |  |  |  |

### Outcome data

#### Acute events frequency among asthmatic versus non-asthmatic controls during the pandemic

Using generalized linear models, we evaluated between group differences in the frequency of acute events during COVID-19 pandemic (table 3). Children with asthma were not found to be at increased risk of LRTIs, episodes of pyrexia, emergency visits or hospitalizations during the pandemic. However, the second model, that accounted for more variables (race, comorbidities, flu vaccination history and passive smoking history), revealed an increased risk of URTI among children with asthma, compared to the control group, during the pandemic (p=0.005).

The strongest predictor for each type of acute events was the number of such events experienced by the participants during the preceding year (p<0.001 for all analyses). In addition, older age was associated with a decreased risk of pyrexia (p=0.025), while the race of the participants was also associated with the frequency of these events.

#### Impact of the pandemic on the frequency of the acute events, compared to the preceding year

Next, using paired statistics, we compared the frequency of each acute event during the pandemic, compared to the preceding year (table 3). Children with asthma experienced fewer URTIs, episodes of pyrexia, emergency visits, hospital admissions, asthma attacks and hospitalizations due to asthma during the pandemic, compared to the preceding year. No differences were observed in the frequency of LRTIs during versus before the pandemic.

On the other hand, the pandemic did not appear to impact the frequency of emergency visits or hospital admissions of non-asthmatic controls. In this group, we observed a decreased frequency of URTIs and episodes of pyrexia, but also a significantly increased risk of LRTIs during the pandemic.

#### Asthma control during the pandemic

We then compared asthma control during versus before the pandemic (table 4). A validated asthma control tool was used, following the local practice in every participating center, to ensure the availability of a measurement of the same tool during the pandemic and one during 2019. At least two asthma control measures (during and before the pandemic) were available for 90.9% of participants in the asthma group. ACT was used in 756 (78.9%) of these children, while cACT, ACQ, or CASI was used in the remaining cases.

**Table 4:** Change in asthma control during the pandemic, compared to 2019. Presented as median [percentiles 25-75. MCID: Minimal Clinically Important Difference. A. Main analysis. B. Sensitivity analysis only including historical asthma control evaluations between March-June 2019.

|  | N | 2019 | Pandemic | % improved or unchanged | % improved | % exceeding MCID |
| --- | --- | --- | --- | --- | --- | --- |
| *a. Main analysis* |  |  |  |  |  |  |
| ACQ<br>(↓=better) | 34 | 0.14 [0 - 0.74] | 0 [0 - 0] | 100% | 56.7% | 26.7%<br>(MCID: 0.5) |
| ACT<br>(↑=better) | 756 | 20 [16 - 23] | 23 [19 - 25] | 89.9% | 67.4% | 32.1%<br>(MCID: 3) |
| cACT (↑=better) | 108 | 22 [19 - 24] | 25 [23 - 26]<br>↑ in 66.7% of subjects | 87.2% | 66.7% |  |
| CASI<br>(↓=better) | 60 | 2 [1 - 5] | 1 [1 - 2] | 93.3% | 50% | 50%<br>(MCID: 0.9) |
| z-scores<br>(↑=better) | 958 |  |  | 90.2% | 65.9% |  |
| *b. Sensitivity analysis only including historical asthma control evaluations between March-June 2019.* |  |  |  |  |  |  |
| ACQ<br>(↓=better) | 15 | 0.14 [0 - 1.01] | 0 [0 - 0] | 100% | 53.3% | 33.3%<br>(MCID: 0.5) |
| ACT<br>(↑=better) | 284 | 22 [18 - 24] | 24 [22 - 25] | 88.7% | 61.6% | 28.5%<br>(MCID: 3) |
| cACT<br>(↑=better) | 11 | 24 [23 - 25] | 25 [23 - 25] | 81.8% | 45.5% |  |
| CASI<br>(↓=better) | 21 | 3 [2 - 5] | 2 [1 - 3] | 95.2% | 57.1% | 57.1% |
| Standardized<br>(↑=better) | 331 |  |  | 89.4% | 60.4% |  |

Improved or unchanged asthma control during pandemic was reported by 90.2% of the participants, while 65.9% experienced an improvement. Importantly, one in three children with asthma (33.2%) reported an improvement in control that exceeded the minimal clinically important difference of the test used.

Given the seasonal differences in asthma control, we conducted a sensitivity analysis where we only included historical asthma control measurements conducted during the same months with the first months of the pandemic (March-June 2019), with consistent findings (table 4).

### Analyses of subgroups

#### Pulmonary function during the pandemic

Pulmonary function during the pandemic compared to 2019 was evaluated in a subgroup of asthma children with available data (table 5). Adequate measurements of pre-bronchodilatation FEV1, post-bronchodilatation FEV1 and PFR were available in 155, 90 and 106 children, respectively. Paired analyses suggested pre-bronchodilatation FEV1 and PFR were significantly improved during the pandemic, while there was also a non-significant trend for improved post-bronchodilatation FEV1.

**Table 5:** Change in pulmonary function during the pandemic, compared to 2019. Presented as mean (SD).

| **Pulmonary function test** | **N** | **Mean 2019** | **Mean pandemic** | **P-value** |
| --- | --- | --- | --- | --- |
| *Main analysis* |  |  |  |  |
| *FEV1 pre-bronchodilatation* | 155 | 94.5% (15.1%) | 98.4% (16.6%) | **<0.001** |
| *FEV1 post-bronchodilatation* | 90 | 100.6% (14.6%) | 104.0% (19.2%) | 0.055 |
| *PFR* | 106 | 121.4% (94.7%) | 129.4% (99.4%) | **0.003** |
| *Sensitivity analysis only including historical asthma control evaluations between March-June 2019.* |  |  |  |  |
| *FEV1 pre-bronchodilatation* | 63 | 93.2% (12.0%) | 97.4% (15.2%) | **0.046** |
| *FEV1 post-bronchodilatation* | 38 | 98.3% (12.8%) | 103.5% (22.5%) | 0.167 |
| *PFR* | 39 | 121.8% (106.5%) | 128.7% (106.6%) | 0.141 |

In a sensitivity analysis only including historical pulmonary function evaluated during the same months with the pandemic (March-June 2019), pre-bronchodilatation FEV1 was significantly improved during the pandemic, while the remaining measures showed non-significant, numerical improvements.

#### Subgroup analysis only including countries that were more severely hit by the pandemic

In this subgroup analysis, all previously described analyses were repeated, including data from participating centers from countries that were more severely hit by the pandemic (>200 deaths per million of inhabitants by 13/07/2020). More specifically, we included data from France, Italy, Mexico, Spain, UK, and the USA. This subgroup analyses, which involved 597 children with asthma and 298 non-asthmatic controls, yielded consistent results with the main analyses (supplementary tables 2-5).

## DISCUSSION

### Key results

In the multinational PeARL childhood asthma cohort, we evaluated the impact of the COVID-19 pandemic on asthma activity. During the pandemic, children with asthma experienced improved disease control (two thirds of the patients), as evidenced by improved scores in validated asthma control measures (>MCID in one third), fewer asthma attacks, fewer hospitalizations, and improved pulmonary function. The multifaceted etiology of this observation may include the avoidance of major asthma triggers including outdoor allergens, viral infections, physical exercise and air pollution, due to social distancing, home sheltering and reduced school days^10,12,22,23,24,25^. Increased treatment adherence during the pandemic^26^, could have also contributed and is consistent with the high adherence levels observed during the pandemic in our cohort. However, along with these ‘protective’ effects of the pandemic, children were also more exposed to indoor allergens and pollutants and possibly adverse psychological factors and this could have precipitated worse asthma control, nevertheless in a minority of children (1/10 children in our cohort).

### Interpretation

The interplay between asthma and COVID-19 infection is being extensively investigated. Several studies explored the prevalence of asthma among children and adults with confirmed COVID-19 infection and failed to find an increased risk of contracting the infection or of adverse outcomes among asthmatic patients^27,28^. A recent review with meta-analysis done in adults with COVID-19 (744 asthmatic and 8151 non-asthmatic) showed that the presence of asthma had no significant effect on mortality (OR=0.96 [0.70-1.30], I^2^=0%, p=0.79), duration of hospitalization, or the risk of ICU admission^29^. However, COVID-19 infection was significantly underdiagnosed during the initial months of the pandemic due to limited availability of diagnostic tests, that led most countries to recommend home isolation without testing in case of non-severe symptoms consistent with COVID-19. The PeARL asthma cohort is the first large, multinational cohort to demonstrate that children with asthma do not experience more frequent events of disease deterioration that could have potentially resulted from increased susceptibility to SARS-CoV-2, not even in countries with an increased COVID-19 burden. In contrast, most outcomes were improved. In addition to suggesting that pediatric asthma is not a risk factor for COVID-19, these findings strongly reinforce the need and potential of compliance to treatment^30,31^, as well as the prospect of improved outcomes with environmental control in asthma, an area of some discrepancy^32^.

The role of different viruses as asthma attack triggers has been investigated extensively. While rhinoviruses represent the main viral trigger of attacks, coronaviruses also trigger asthma attacks, albeit less frequently^33^. However, and in line with our findings, the outbreak of SARS in Singapore and Hong Kong were not associated with increased asthma attacks in children^34,35^. On the contrary, and in line with our results, the incidence of acute respiratory tract infections and acute asthma attacks declined dramatically, likely due to the closure of schools for a period of time, stepped-up public hygiene measures and the use of facemasks^36,37^.

On the other hand, improved outcomes among asthmatic children during SARS/COVID-19 epidemics may also support the hypothesis of a protective effect of asthma against COVID-19. This hypothesis is based on several observations, including changed in the immune response or decreased risk secondary to chronic medications such as inhaled corticosteroids (ICS). In-vitro models have shown that ICS may suppress both coronavirus replication and cytokine production^38,39^. Analysis of induced sputum samples in a well-characterized cohort of adults with severe asthma found reduced ACE2 (angiotensin-converting enzyme 2) and TMPRSS2 (transmembrane protease serine 2) gene expression among patients taking ICS, and especially among those on higher doses^40^; and we know ACE2 and TMPRSS2 mediate SARS-CoV-2 cell infection. Similarly, a recent study done in children and adults showed that patients with asthma and respiratory allergies had reduced ACE2 gene expression in airway cells, suggesting a potential mechanism of reduced COVID-19 risk^11^.

### Limitations and strengths

Evaluation of the frequency of acute events during the pandemic is challenging, but we believe our methodology was rigorous. Firstly, the accuracy of parent-reported frequency of acute events has been previously validated^41^. We assumed a potential under-reporting of acute events during 2019, due to recall bias, however we do not anticipate between group differences in this bias. In addition, potential under-reporting of acute events prior to the pandemic would have led to an overestimation of the relative risk during the pandemic. Since we did not find an increased risk, we suggest the impact of recall bias was minimal.

For estimating the duration of the pandemic, we considered the onset of the pandemic to coincide with the first fatality in each respective country. This usually followed a few days to weeks after the identification of the first case. We chose this time-point as an objective marker that could better identify the onset of the impact of the pandemic in each community. Since we were inquiring for acute episodes during the epidemic, we wanted to avoid a potential underestimation of the frequency of the events that could have been caused by accounting for a wider time-period of the epidemic.

Extrapolating an annualized asthma attack rate based on the events that were observed during the pandemic is complicated by the seasonal variability of asthma attacks^42,43^. However, numerous high-quality studies demonstrate a peak in childhood asthma attack during autumn and schools reopening, which is counter-balanced by a low frequency during summer^42,44^. Interestingly, these studies consistently show that the frequency of asthma attacks and asthma hospitalizations during spring and early summer levels with the median frequency of asthma attacks throughout the year^42,43^. The study period coincided with these months, suggesting that the frequency of acute events equals to their frequency throughout the year. This observation enhances our confidence on the accuracy of our annualize estimates and our findings, however the limited observation period during the pandemic remains a limitation of this study.

The over-representation of atopic diseases in the control group shall be mentioned as a potential limitation, but it has been accounted for in our analyses. Finally, different validated questionnaires were used for assessing asthma control across the study centres, depending on the availability of historic measurements, to allow for paired comparisons. While the results of different tools are not directly comparable, our study revealed consistent results in the subgroups evaluated with different tools. On the other hand, the extensive, multinational study population with a good geographic balance across 4 continents, was a major strength of our study, increasing our confidence in our findings.

## Conclusion

Overall, this analysis of the PeARL childhood asthma cohort revealed improved health and asthma activity during the COVID-19 pandemic, probably associated with decreased exposure to asthma triggers and increased treatment adherence. It also demonstrated that during the pandemic, children with predominantly mild to moderate atopic asthma did not suffer from an increase in the frequency of acute episodes that could represent COVID-19 infection, refuting the hypothesis that childhood asthma is a risk factor for COVID-19.

## Data Availability

Deidentified patient level data will be available upon request between 3 months and 5 years after the publication of this manuscript.

## Acknowledgements

This study was supported by the Respiratory Effectiveness Group (REG). REG has received support from AstraZeneca, Novartis and Sanofi for continued work on PeARL. AGM was supported by the National Institute of Health Research Manchester Biomedical Research Centre (NIHR Manchester BRC). We thank Mrs Maria Kritikou for excellent administrative support of the study.

## Online supplement

### PeARL Steering Committee

#### Principal Investigator

*Prof. Nikolaos G. Papadopoulos*, University of Athens, Greece & University of Manchester, UK.

#### Members

*Prof. Adnan Custovic*, Imperial College London, UK.

*Prof. Antoine Deschildre*, Hôspital Jeanne de Flandre, CHU Lille, France.

*Dr. Alexander G. Mathioudakis*, University of Manchester, UK.

*Prof. Wanda Phipatanakul*, Children’s Hospital Boston, USA.

*Prof. Gary Wong*, The Chinese University of Hong Kong, Hong Kong.

*Prof. Paraskevi Xepapadaki*, University of Athens, Greece.

## Supplementary tables

**Supplementary Table 1:** Country / continent distribution of the responses.

| Country | Children with asthma | Healthy controls | Number of centres |
| --- | --- | --- | --- |
| **Africa** |  |  |  |
| Egypt | 90 (8.7%) | 50 (10.1) | 2 |
| **Americas** |  |  |  |
| Mexico | 184 (17.5%) | 90 (17.8%) | 4 |
| USA | 60 (5.7%) | 30 (5.9%) | 1 |
| **Asia** |  |  |  |
| China | 67 (6.4%) | 32 (6.3%) | 1 |
| Hong Kong | 30 (2.9%) | 15 (3.0%) | 1 |
| India | 24 (2.3%) | 12 (2.4%) | 1 |
| **Europe** |  |  |  |
| Finland | 60 (5.7%) | 30 (5.9%) | 1 |
| France | 190 (18.0%) | 86 (17.0%) | 6 |
| Greece | 34 (3.2%) | 16 (3.2%) | 1 |
| Italy | 32 (3.0%) | 15 (3.0%) | 1 |
| Poland | 62 (5.9%) | 31 (6.1%) | 1 |
| Romania | 30 (2.9%) | 11 (2.2%) | 1 |
| Russia | 60 (5.7%) | 30 (5.9%) | 1 |
| Spain | 109 (10.3%) | 55 (10.9%) | 2 |
| UK | 22 (2.1%) | 2 (0.4%) | 1 |

**Supplementary table 2:**
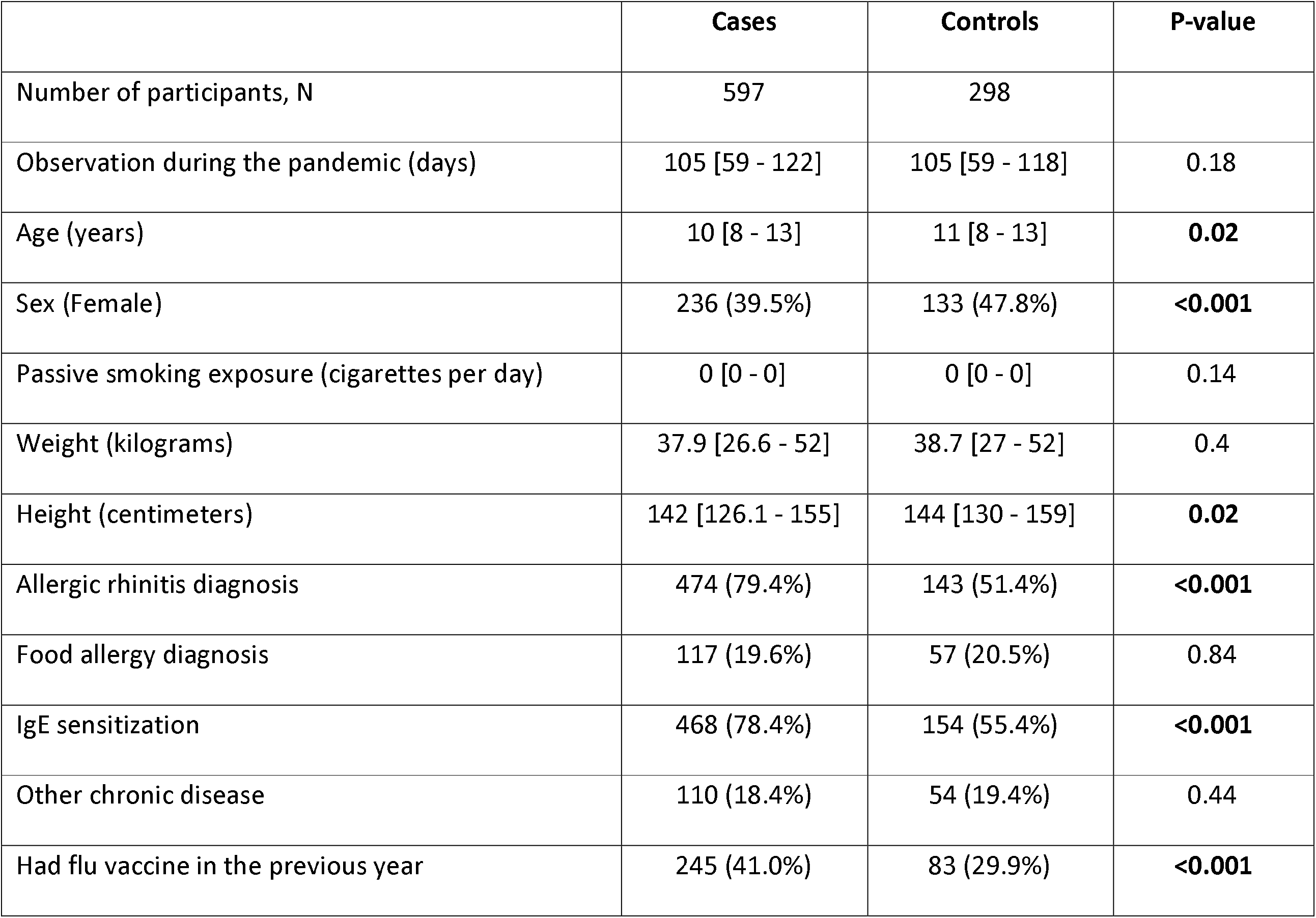
Main characteristics of the participants <u>from countries with high COVID-19 burden</u>. Continuous data are presented as median [percentiles 25-75], total number of participants with available data for this variable. and dichotomous data as number of events/ total numbers (percentage).

|  | Cases | Controls | P-value |
| --- | --- | --- | --- |
| Number of participants, N | 597 | 298 |  |
| Observation during the pandemic (days) | 105 [59 - 122] | 105 [59 - 118] | 0.18 |
| Age (years) | 10 [8 - 13] | 11 [8 - 13] | **0.02** |
| Sex (Female) | 236 (39.5%) | 133 (47.8%) | **<0.001** |
| Passive smoking exposure (cigarettes per day) | 0 [0 - 0] | 0 [0 - 0] | 0.14 |
| Weight (kilograms) | 37.9 [26.6 - 52] | 38.7 [27 - 52] | 0.4 |
| Height (centimeters) | 142 [126.1 - 155] | 144 [130 - 159] | **0.02** |
| Allergic rhinitis diagnosis | 474 (79.4%) | 143 (51.4%) | **<0.001** |
| Food allergy diagnosis | 117 (19.6%) | 57 (20.5%) | 0.84 |
| IgE sensitization | 468 (78.4%) | 154 (55.4%) | **<0.001** |
| Other chronic disease | 110 (18.4%) | 54 (19.4%) | 0.44 |
| Had flu vaccine in the previous year | 245 (41.0%) | 83 (29.9%) | **<0.001** |

**Supplementary table 3:** Asthma severity and asthma treatment adherence in the subgroup of children with asthma <u>from countries with high COVID-19 burden</u>.

|  |  |
| --- | --- |
| **Asthma severity:**<br>- Intermittent<br>- Mild persistent<br>- Moderate persistent<br>- Severe persistent<br>- Unknown | 190 (31.8%)<br>189 (31.7%)<br>146 (24.5%)<br>67 (11.2%)<br>5 (0.84%) |
| **Treatment adherence:**<br>- Always as prescribed<br>- Sometimes not taken<br>- Often not taken<br>- Never taken<br>- Unknown | 310 (51.9%)<br>137 (23.0%)<br>33 (5.5%)<br>70 (11.7%)<br>47 (7.9%) |

**Supplementary table 4:**
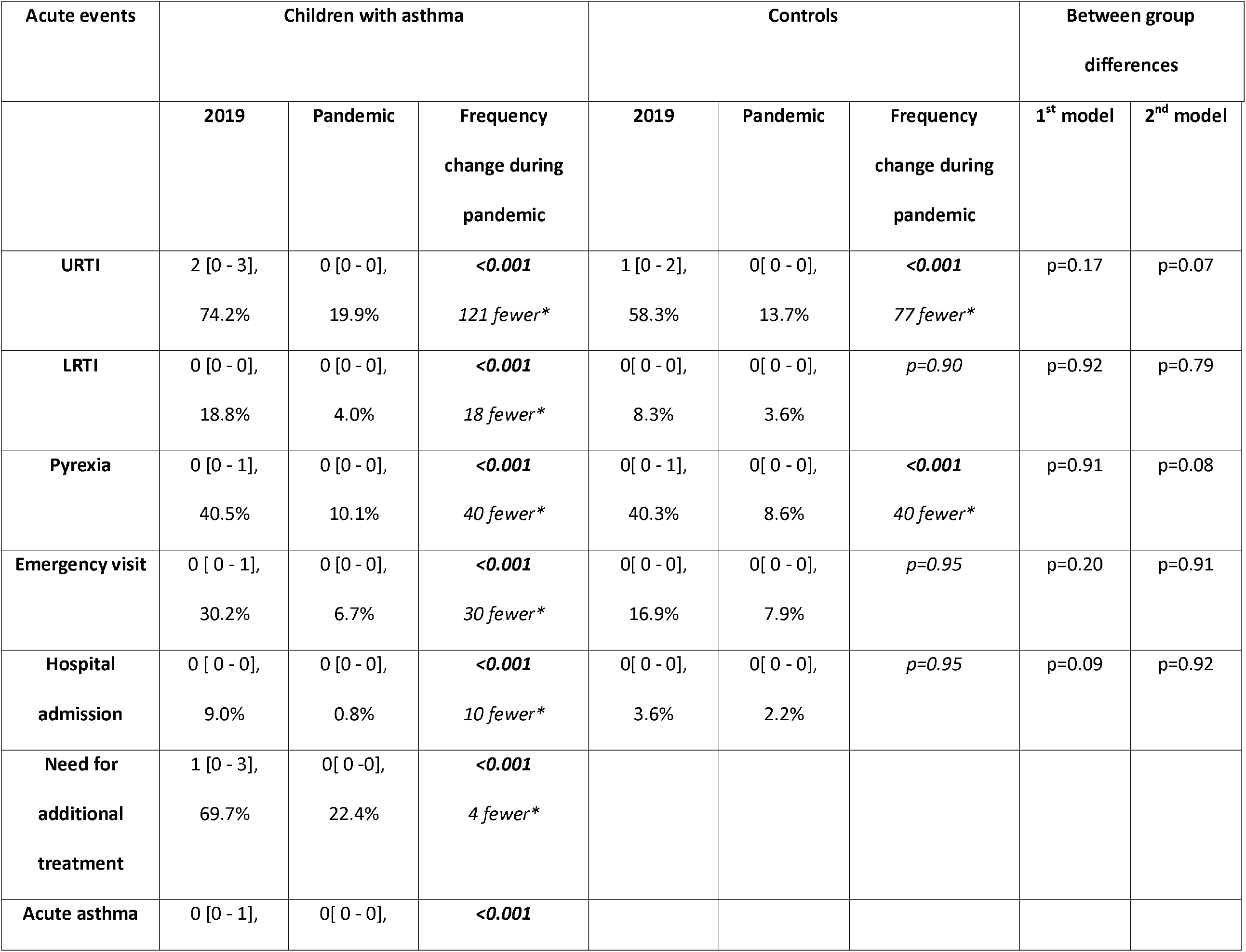
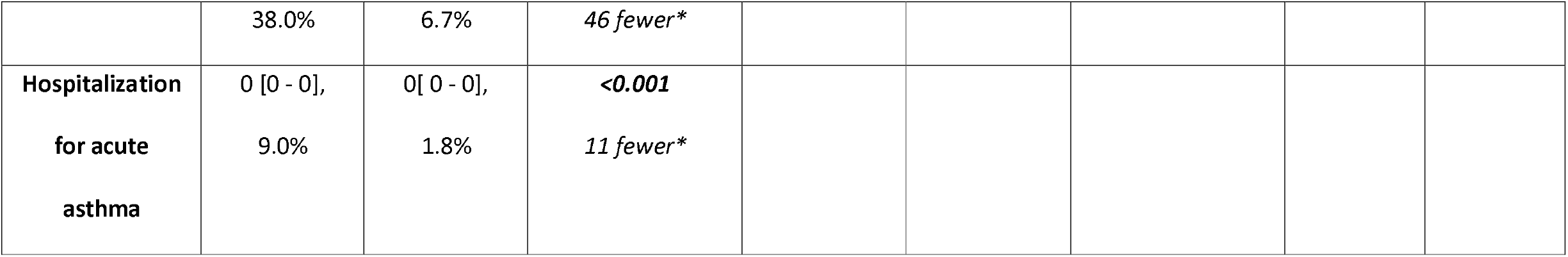
Acute events observed during 2019 and during the pandemic participants <u>in the subgroup of participants from countries with high COVID-19 burden</u>. Presented as median [percentiles 25-75], and the percentage of participants experiencing at least one event in the respective observation period. <u>* Events per 100 participants per year.</u>

**Supplementary table 5:** Change in asthma control during the pandemic, compared to 2019 <u>in the subgroup of participants from countries with high COVID-19 burden</u>. Presented as median [percentiles 25-75]. MCID: Minimal Clinically Important Difference.

|  | N | 2019 | Pandemic | % improved or unchanged | % improved | % exceeding MCID | p |
| --- | --- | --- | --- | --- | --- | --- | --- |
| *Main analysis* |  |  |  |  |  |  |  |
| ACQ (↓better) | 0 |  |  |  |  |  |  |
| ACT (↑better) | 430 | 21 [18 - 24] | 24 [22 - 25] | 90.2% | 64.6% | 36%<br>(MCID: 3) | **<0.001** |
| cACT (↑better) | 0 |  |  |  |  |  |  |
| CASI (↓better) | 60 | 2 [1 - 5] | 1 [1 - 2] | 93.3% | 50% | 50%<br>(MCID: 0.9) | **<0.001** |
| z-scores<br>(↑better) | 490 |  |  | 90.6% | 62.9% |  | **<0.001** |
| *Sensitivity analysis only including historical asthma control evaluations between March-June 2019.* |  |  |  |  |  |  |  |
| ACQ (↓better) | 0 |  |  |  |  |  |  |
| ACT (↑better) | 207 | 22 [18 - 24] | 24 [22 - 25] | 88.9% | 62.8% | 29.0%<br>(MCID: 3) | **<0.001** |
| cACT (↑better) | 0 |  |  |  |  |  |  |
| CASI (↓better) | 21 | 3 [2 - 5] | 2 [1 - 3] | 95.2% | 57.1% | 57.1% | **<0.001** |
| Standardized<br>(↑better) | 228 |  |  | 89.5% | 62.3% |  | **<0.01** |

**Supplementary table 6:** Change in pulmonary function during the pandemic, compared to 2019 <u>in the subgroup of participants from countries with high COVID-19 burden</u>. Presented as mean (SD).

| ***Pulmonary function test*** | ***N*** | ***Mean 2019*** | ***Mean pandemic*** | ***P-value*** |
| --- | --- | --- | --- | --- |
| *FEV1 pre-bronchodilatation* | 84 | 94.8% (14.85%) | 97.9% (17.9%) | 0.04 |
| *FEV1 post-bronchodilatation* | 70 | 101.0% (14.1%) | 103.4% (20.3%) | 0.25 (NS) |
| *PFR* | 72 | 135.9% (108.2%) | 142.6% (113.2%) | 0.06 (NS) |

